# Uncertainty Quantification in COVID-19 Detection Using Evidential Deep Learning

**DOI:** 10.1101/2022.05.29.22275732

**Authors:** Bardia Khosravi, Shahriar Faghani, Amir Ashraf-Ganjouei

## Abstract

Considering the immense pace of developments in deep learning (DL), its applications in medicine are relatively limited. One main issue that hinders the utilization of DL in the medical practice workflow is its reliability. A radiologist interpreting an image can easily say “I don’t know”, while a DL model is forced to output a result. Evidential deep learning (EDL) is one of the methods for uncertainty quantification (UQ). In this work, we aimed to use EDL to express model uncertainty in detecting COVID-19. We used SIIM-FISABIO-RSNA COVID-19 chest x-ray dataset and trained a model to diagnose typical COVID-19 pneumonia. When applied to a separate test set, it yielded an accuracy of 88% with median uncertainty scores of 0.25 and 0.07 for normal and typical COVID-19 images, respectively. Moreover, the model labeled unseen indeterminate and atypical COVID-19 x-rays with median uncertainties of 0.32 and 0.35, respectively. Our model’s performance was superior to the exact model trained with conventional approach of DL (i.e., using the cross-entropy loss), which is not able to express the uncertainty level. Overall, this study demonstrates applicability of UQ in disease detection that could facilitate the use of DL in practice by increasing its reliability.

## Introduction

Artificial intelligence and deep learning (DL) frameworks can assist physicians by automating image analysis in terms of detection of pathological findings, characterization of pathologies and quantification of disease extent [1, 2]. However, it is remarkably challenging to deploy these tools in clinical settings even for simple tasks such as analyzing chest x-ray (CXR) images. One of the factors that makes them less applicable is the presence of performance errors. Therefore, most studies in recent years have focused on implementing novel methods to improve their accuracy. However, current models lack the implicit ability to say *“I don’t know”* when provided input data that is out-of-distribution for the training data; compared to radiologists who tend to be careful when unsure about the diagnosis [3]. Therefore, there has been a recent interest in measuring the uncertainty level of DL models [4].

Uncertainty quantification aims at characterizing the model’s predictions with a confidence measure, which can be obtained through various approaches [4]. For instance, Sensoy et al. described evidential deep learning (EDL) for uncertainty quantification and tackled this problem from the perspective of *Theory of Evidence* [5]. By replacing the softmax function, which is considered as the standard output of a classification network, with the parameters of a Dirichlet density distribution, the model learns to recognize data instances that contribute efficiently to the performance during training while providing an uncertainty estimate. This EDL model has a specific loss function, such that it can be minimized using backpropagation.

Starting in late 2019, the COVID-19 virus rapidly dominated the entire world, causing global pandemic and economic crises. Although not recommended as a screening tool, CXR can be a valuable modality when it comes to COVID-19 diagnosis, especially in resource-scarce centers. Since COVID-19 infection can present with atypical presentations on CXR, we aimed to quantify the uncertainty estimates of a deep learning model for diagnosing COVID-19 infection. To the best of our knowledge, this is the first study implementing evidential deep learning to measure uncertainty in COVID-19 diagnosis.

## Material and Method

### Dataset

In this study, we used the SIIM-FISABIO-RSNA COVID-19 chest x-ray dataset which consists of 6054 de-identified studies (both anteroposterior and posteroanterior) categorized into four groups based on presence of signs of COVID-19 pneumonia: negative for pneumonia (no lung opacities; n=1676), typical appearance (multifocal bilateral, peripheral opacities with rounded morphology, lower lung–predominant distribution; n=2855), indeterminate appearance (absence of typical findings and unilateral, central or upper lung predominant distribution; n=1049) and atypical appearance (pneumothorax, pleural effusion, pulmonary edema, lobar consolidation, solitary lung nodule or mass, diffuse tiny nodules or cavity; n=474) [6]. The data was divided into train (90%) and test (10%) sets while ensuring same ratio of the mentioned groups in both sets. Furthermore, we removed duplicate images for each study and normalized the data to have a zero mean and unit standard deviation. We augmented our data by randomly changing intensity (± 0.2), moving (± 10% in each plane) and rotating (± 15°) the images using the MONAI framework [7]. We used a P-100 GPU for all training instances.

### Evidential Deep Learning

The EDL model tries to collect as much evidence on the training task and utilizing the Dempster–Shafer Theory of Evidence (DST) calculates the level of uncertainty for each sample [5]. For a *K* class classification task, DST assigns a belief mass (*b*_*k*_) for each mutually exclusive class (*k*=1, …*K*) and also provide an overall uncertainty mass (*u*) that sum up to one:

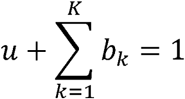

Considering evidence collected for each class (*e*_*k*_) as the non-negative output of the last linear layer of the model corresponding to class *k*, the belief mass for class *k*(*b*_*k*_) is:

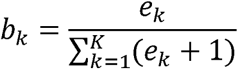

then uncertainty (*u*) would be:

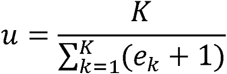

So, when the output of the last linear layer of the model is close to zero (i.e., having collected a few evidences) the model is highly uncertain. We can use {*e*_*k*_+1|*k* ∈ 1,…, *K*}as parameters of the Dirichlet distribution and use Kullback–Leibler divergence (KL) loss to force the model to be uncertain especially when its prediction is incorrect. The latter can be enforced using a second loss functions such as mean squared error, log likelihood or digamma [5].

### Training and Evaluation Parameters

We ran our training pipeline with a variety of models and hyper parameters, list of which are presented in Table-1. Here we go over the final selected model and hyperparameters based on the lowest cross validation loss of five random folds in our experiments. We used the PyTorch implementation of EfficientNet B0 network to do the feature extraction [8, 9]. We used transfer learning from pre-trained weights on natural images so that our model converges faster. We used a classifier head with two linear layers converting 1280 extracted features to 500 and then to the two classes. We combined log-likelihood loss with the KL loss to maximize model uncertainty in the setting of incorrect classification. To prevent erratic increase of the KL loss we used an annealing coefficient of 10, meaning during the first 10 epochs the effect of KL loss increases linearly in each training step. We used the MADGRAD as our optimizer as it outperformed other tested alternatives [10].

**Table 1.** Training grid-search parameters.

|  |  |  |  |  |  |
| --- | --- | --- | --- | --- | --- |
| Model | EfficientNet B0<sup>‡</sup> | Batch Size | 8 | Learning Rate | 5e-3 |
|  | EfficientNet B1 |  | 32<sup>‡</sup> |  | 1.25e-3<sup>‡</sup> |
|  | ResNet 50 |  | 64 |  | 6.25e-4 |
|  | RegNetY 400 |  | 256 |  | 6.25e-5 |
| Optimizer | Adam | Learning Rate Scheduler | Step<sup>‡</sup> | Image Size | 224 x 224<sup>‡</sup> |
|  | SGD |  | OneCycle |  | 380 x 380 |
|  | MADGRAD<sup>‡</sup> |  | No Scheduler |  | 512 x 512 |

The parameters used in our preliminary grid-search. The option selected for our final configuration is denoted with <sup>‡</sup>.

To measure the model’s performance both in terms of accuracy and uncertainty on seen and unseen categories, we first removed all atypical and indeterminate samples from our training and test sets, and trained the model on normal images and typical COVID-19 pneumonia studies as a binary classification task with 1:1 class ratio. Moreover, we inferred uncertainty estimates on two distinct sets of atypical and indeterminate CXRs. We also trained an exact model with the same architecture and hyperparameters but with the cross-entropy loss to compare the loss functions’ convergence capabilities.

## Results

Our final dataset comprised of 3373 male participants (55.71%). After training the EDL model, it reached a final accuracy of 88.77%, though training the same model with the cross-entropy loss yielded an accuracy of 87.22% over the test set. On the test set, the EDL model provided median uncertainty score of 0.25 and 0.07 for normal and typical COVID-19 CXRs, respectively. Figure-1 demonstrates that when the accuracy declines, the EDL model provides higher uncertainty scores and as depicted in Figure-2, the EDL model was able to provide a wide range of uncertainty scores for CXRs with typical COIVD-19 infection. Moreover, we tested our model on the indeterminate and atypical subsets and the median uncertainty scores were 0.32 and 0.35, respectively. Figure-3 illustrates examples from normal, indeterminate and atypical subsets.

**Figure 1.**
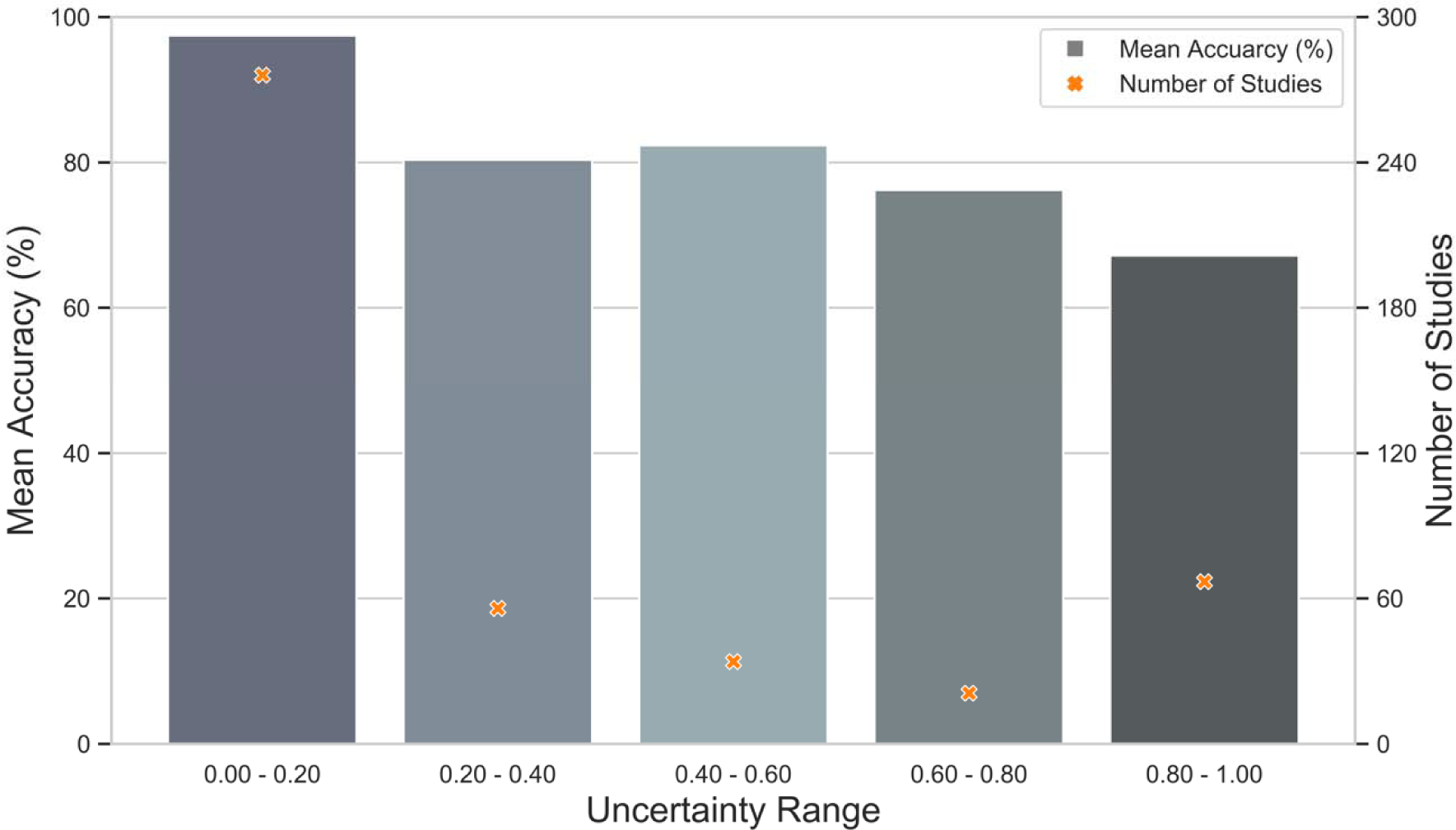
EDL model accuracy based on estimated uncertainty scores. Mean accuracy of the test subset categorized based on the estimated uncertainty score.

**Figure 2.**
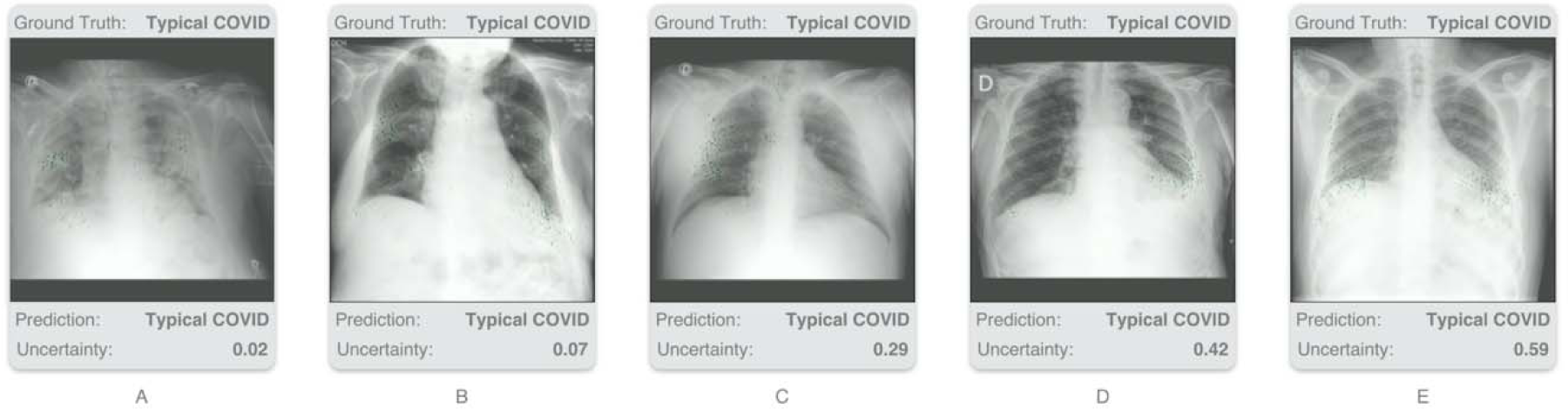
CXRs with COVID-19 infection. This figure exemplifies five lung suffering from COVID-19 infection. A typical COVID-19 CXR was given an uncertainty score of 0.02 (A), whereas an uncharacteristic presentation had an uncertainty score of 0.59 (E).

**Figure 3.**
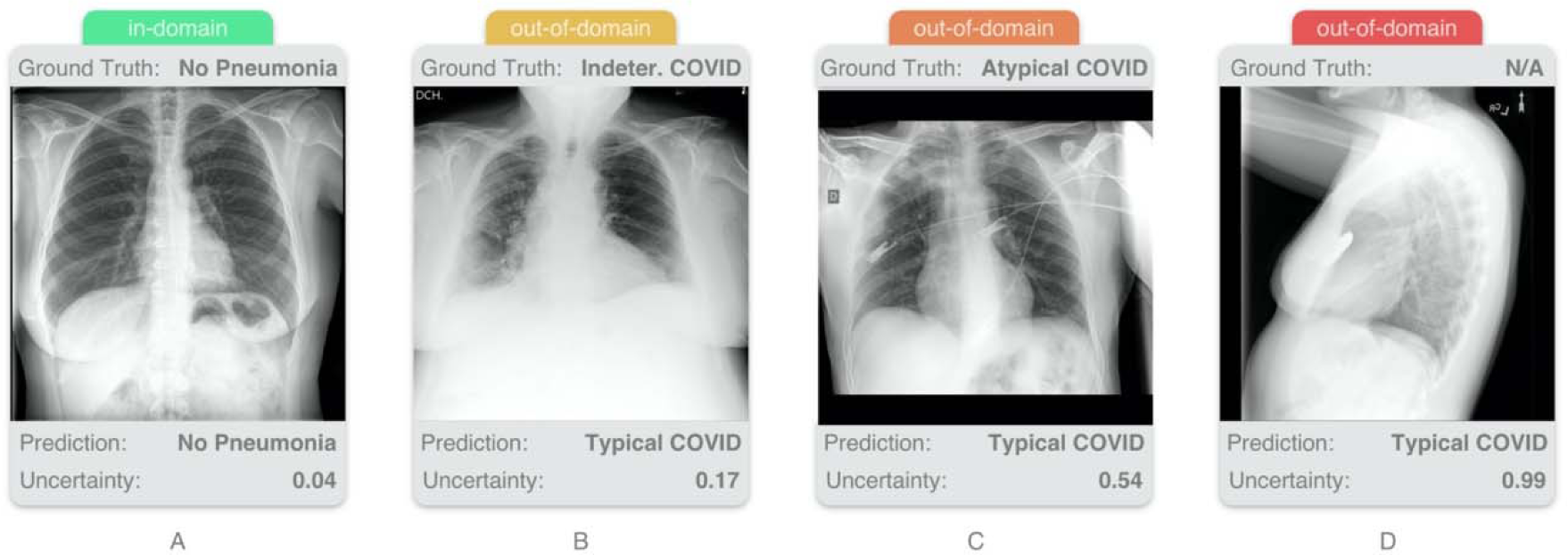
CXRs from normal, indeterminate and atypical subsets. As depicted here, a normal CXR with uncertainty score of 0.04 (A), whereas giving a lateral normal CXR to the EDL model yielded an uncertainty score of 0.99 (D). B and C illustrate examples from indeterminate and atypical subsets with uncertainty scores of 0.17 and 0.54, respectively.

## Discussion

Despite the fact that DL frameworks have achieved outstanding results in medical imaging tasks (specially classification), majority of these models are overconfident and have not been deployed due to their lack of reliability, risk of erroneous decisions and poor generalization on unseen data [4, 11]. In the context of COVID-19 CXR classification problems, we have implemented a method that is capable of differentiating COVID-19 pneumonia while also reporting certainty of its predictions. We have demonstrated that the EDL model has an accuracy as high as the conventional models, and it is able to provide higher uncertainty scores in cases with uncharacteristic and indeterminate imaging presentations of COVID-19 pneumonia in comparison with typical cases.

In recent years, various uncertainty quantification methods have been proposed in the literature, including ensemble approaches, Bayesian methods, etc. [12]. For instance, Tardy and colleagues combined two uncertainty measurements to reject outliers in a mammogram classification task [13]. Furthermore, several approaches have been suggested based on a Bayesian formulation of the uncertainty. Although Bayesian approaches are highly complex, multiple frameworks have been proposed based on the Bayesian formulation of the uncertainty because these networks are capable of isolating different sources of uncertainty. In this regard, multiple medical imaging studies have used Bayesian DL methods such as Monte Carlo Dropout, variational inference and variational auto encoders [14-16].

Following a Bayesian approach, EDL framework models the output of the network with a Dirichlet distribution [5]. We demonstrated that by using an uncertainty-aware loss function in the EDL framework, the system learns not only the probabilistic estimate for classification, but also estimates the uncertainty of its predictions. It was shown that this method can out-perform other uncertainty quantification methods in terms of accuracy, training speed and uncertainty estimation on natural image datasets [5, 17]. Hemmer and colleagues demonstrated that EDL can be successfully applied to a medical imaging classification task [14]. Although their results were promising, their smaller dataset hindered their model’s generalization capabilities.

## Conclusions

In conclusion, the current work at hand provides an insight into the effective implementation of an uncertainty quantifying method for DL-assisted diagnosis of COVID-19. The current method has several strengths such as a high accuracy of prediction, fast training speed and measuring uncertainty scores from only a single run. However, a shortcoming of this approach may be the absence of an uncertainty estimate cut-off to indicate studies requiring second opinion from an expert radiologist. We might be able to rely on deep learning models to measure uncertainty in future, though more research is required to address the limitations in this regard.

## Data Availability

All data produced are available online at:
https://www.kaggle.com/competitions/siim-covid19-detection/data

https://www.kaggle.com/competitions/siim-covid19-detection/data

## Acknowledgments

None.

